# Metabolome-wide association study on *ABCA7* demonstrates a role for ceramide metabolism in impaired cognitive performance and Alzheimer’s disease

**DOI:** 10.1101/2021.04.06.21254991

**Authors:** Abbas Dehghan, Rui Pinto, Ibrahim Karaman, Jian Huang, Brenan R Durainayagam, Sonia Liggi, Luke Whiley, Rima Mustafa, Miia Kivipelto, Alina Solomon, Tiia Ngandu, Takahisa Kanekiyo, Tomonori Aikawa, Elena Chekmeneva, Stephane Camuzeaux, Matthew R. Lewis, Manuja R Kaluarachchi, Mohsen Ghanbari, M Arfan Ikram, Elaine Holmes, Ioanna Tzoulaki, Paul M. Matthews, Julian L. Griffin, Paul Elliott

**Author notes:** **Corresponding author:** Professor Paul Elliott, MRC-HPA Centre for Environment and Health, Department of Epidemiology and Biostatistics, School of Public Health, Imperial College of Science Technology and Medicine, London W2 1PG, United Kingdom, Abbas Dehghan MD PhD, Department of Epidemiology & Biostatistics, School of Public Health, Imperial College of Science Technology and Medicine, London W2 1PG, United Kingdom. **Competing interests** PMM has received consultancy fees from Biogen, Roche, Celgene and Ipsen Pharmaceuticals. He has received honoraria or speakers’ fees from Novartis, Biogen, Neurodiem and Medscape and has received research or educational funds from Biogen, Novartis and GlaxoSmithKline.

## Abstract

Genome-wide association studies (GWAS) have identified genetic loci associated with risk of Alzheimer’s disease (AD), but underlying mechanisms are largely unknown. We conducted a metabolome-wide association study (MWAS) of AD-associated loci from GWAS using untargeted metabolic profiling (metabolomics) by ultra-performance liquid chromatography-mass spectrometry (UPLC-MS). We identified an association of lactosylceramides (LacCer) with AD-related single nucleotide polymorphisms (SNPs) in *ABCA7* (*P* = 5.0x 10^−5^ to 1.3 x 10^−44^). We show that plasma LacCer concentrations are associated with cognitive performance in humans and concentrations of sphingomyelins, ceramides, and hexose-ceramides were altered in brain tissue from *ABCA7* knock out mice, compared to wild type (WT) (*P* =0.049 to 1.4 x10^−5^). We then used Mendelian randomisation to show that the association of LacCer with AD risk is potentially causal. Our work suggests that risk for AD arising from functional variations in *ABCA7* are mediated at least in part through ceramides. Modulation of their metabolism or downstream signalling may offer new therapeutic opportunities for AD.

---

In recent years, genome-wide association studies (GWAS) have robustly identified thousands of genetic loci for complex traits and disorders. Genetic loci have been used to identify pathways affected in early stages of disease and to differentiate potential causal pathways from pathological processes secondary to their chronic progression in diseases such as Alzheimer’s disease (AD). GWAS have so far identified 47 unique genetic loci for AD^1-3^. Mechanistic understanding of familial-early onset AD arising from variants in genes for amyloid precursor protein (*APP*), presenilin 1 (*PSEN1*) or presenilin 2 (*PSEN2*) has centred around the processing of *APP*, but for most other identified loci contributing to the much more common, sporadic, late onset form of the disease, the underlying pathway(s) mediating risk are less well understood or unknown.

Untargeted metabolomics allows the identification of metabolites and other small molecules in biological samples and provides an objective means to investigate the metabolic consequences of differences in genetic background, endogenous physiological processes and exogenous (environmental) exposures on disease risk{Lotta, 2016 #10;Wittemans, 2019 #11;Tzoulaki, 2019 #13;Holmes, 2008 #14;Illig, 2010 #15}. For AD, metabolomics has been applied most commonly to patient cohorts in small, underpowered studies with typically fewer than 200 cases. Recently, larger studies have found associations between abnormalities in bile acids and AD or mild cognitive impairment (MCI)^9^ and an association between d18:1/24:1 dihexosyl-ceramide and *ABCA7* was reported based on study of 1954 ethnically Chinese individuals^10^.

We conducted a metabolome-wide genetic association study (MWAS) for two epidemiologic cohorts – the Airwave Health Monitoring Study (Airwave)^11^ and the Rotterdam Study (RS)^12^ using ultra-performance liquid chromatography-mass spectrometry (UPLC-MS) and genetic variants identified in prior AD GWAS studies^1-3^. We then tested for associations of the identified metabolites with cognitive measures in Airwave and for participants in the Finnish Geriatric Intervention Study to Prevent Cognitive Impairment and Disability (FINGER)^13^, a clinical trial of lifestyle interventions to prevent or delay dementia among an elderly cohort. We also performed a target lipidomic profile of ceramides and metabolically related molecules in brain tissue from the *Abca7* knockout (KO) mouse^14^. Finally, we applied Mendelian randomisation^15^ to assess potential causal roles of the identified metabolites in determining later life cognitive decline and the risk of AD.

## Results

The study workflow is summarised in **Supplementary Figure 1**.

### MWAS on AD-related SNPs identifies associations between ABCA7 and LacCers

In our UPLC-MS analysis of the Airwave and Rotterdam Study data, we measured 5199 mass spectral features (the unique mass to charge ratio-retention time pairing, m/z-RT) and annotated 1815 of them (**Supplementary Table 1**). The 47 AD-related SNPs that were associated with metabolic features at a metabolome-wide significance level (MWSL, see **Supplementary Methods**) are shown in the Manhattan plot in **Figure 1**. We observed the strongest association (Pearson correlation coefficient = -0.53, *P* = 7.16 x 10^−44^) between rs3752246 in *ABCA7* and a feature with *m/z* = 1068.696 and RT = 7.84 min in the negative ionisation mode spectrum (abbreviated as LNEG, 1068.6958_7.8401) which corresponded to LacCer (d18:1/24:1) (**Figure 1, Supplementary Table 2**). Overall, we identified 121 associations with 10 AD-related SNPs at *ABCA7* that surpassed the significance threshold **(Supplementary Table 2)**. These features corresponded to 41 unique mass spectrometry features (MWSL α = 0.05) mapped to 16 metabolites. (**Figure 2, Supplementary Table 2, Supplementary Methods**). Ten of the 41 metabolic features were from LacCer or sulfatide hexosyl ceramide (SHexCer) which were also associated with two AD-related SNPs in *APOE*, suggesting common metabolic pathways underlying AD risk for *ABCA7* and *APOE*. Using expression quantitative trait loci (eQTL) data from The Genotype-Tissue Expression (GTEx) project^16^, we found that the 10 *ABCA7* SNPs were associated with lower expression of *ABCA7* in multiple human brain tissues including the cerebellum and cerebral hemisphere (**Supplementary Figure 2**). Given the specificity and strength of the metabolite associations for the *ABCA7* locus, we focused the remainder of our study on defining the pathways linking *ABCA7* and risk of AD.

**Figure 1.**
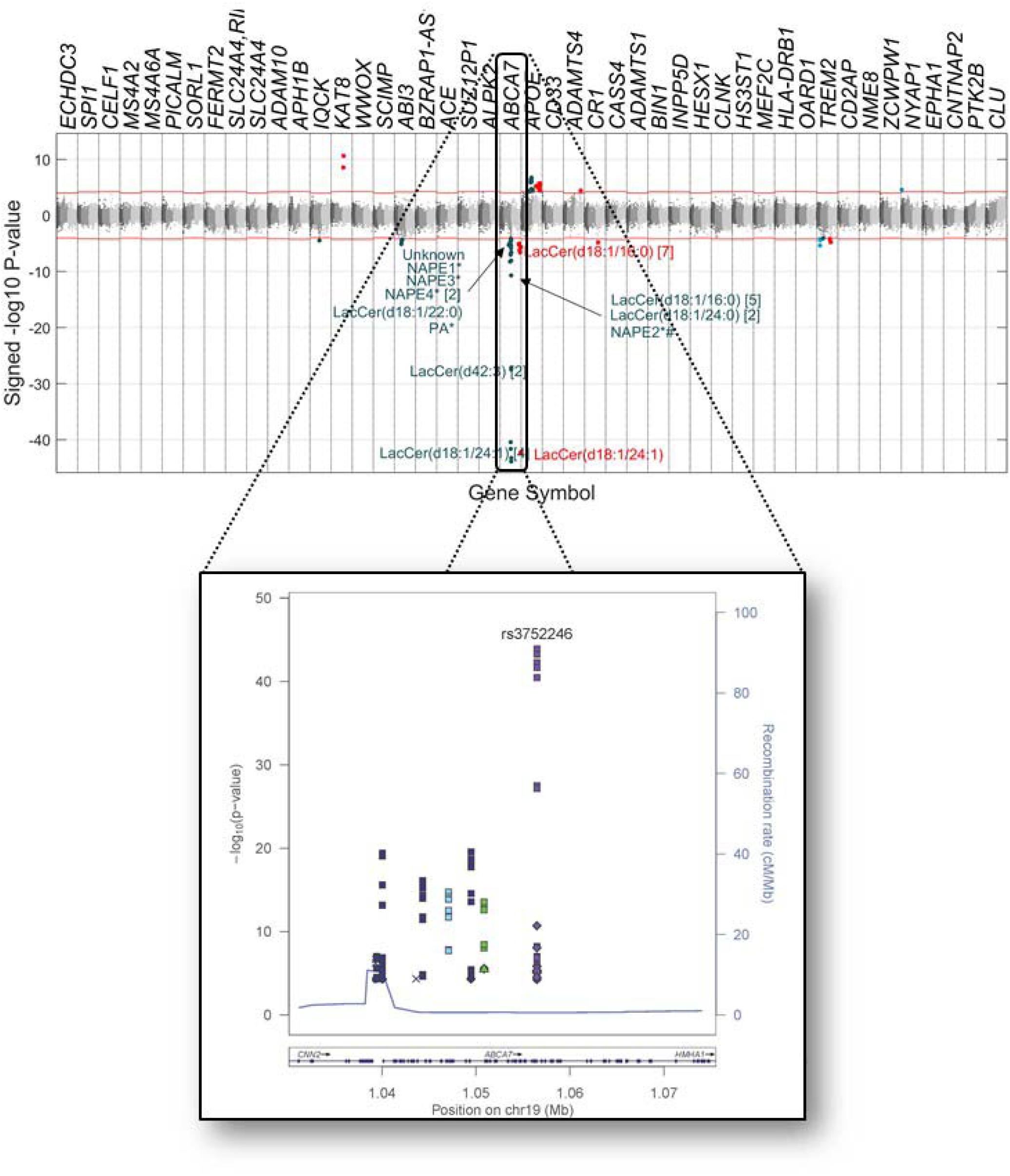
Manhattan plot on the association of metabolites detected by mass spectrometry with AD-related SNPs. Features associated with *ABCA7* SNPs are annotated. For some metabolites more than one metabolic feature was found to be associated with AD-related SNPs and the number of metabolic features corresponding to the same metabolites are described in brackets. Key: LacCer: lactosylceramide, NAPE: N-acyl phosphatidylethanolamine, PA: phosphatidic acid. Red features are assayed by negative ionization mode lipidomics and blue features by positive ionization mode lipidomics.

**Figure 2.**
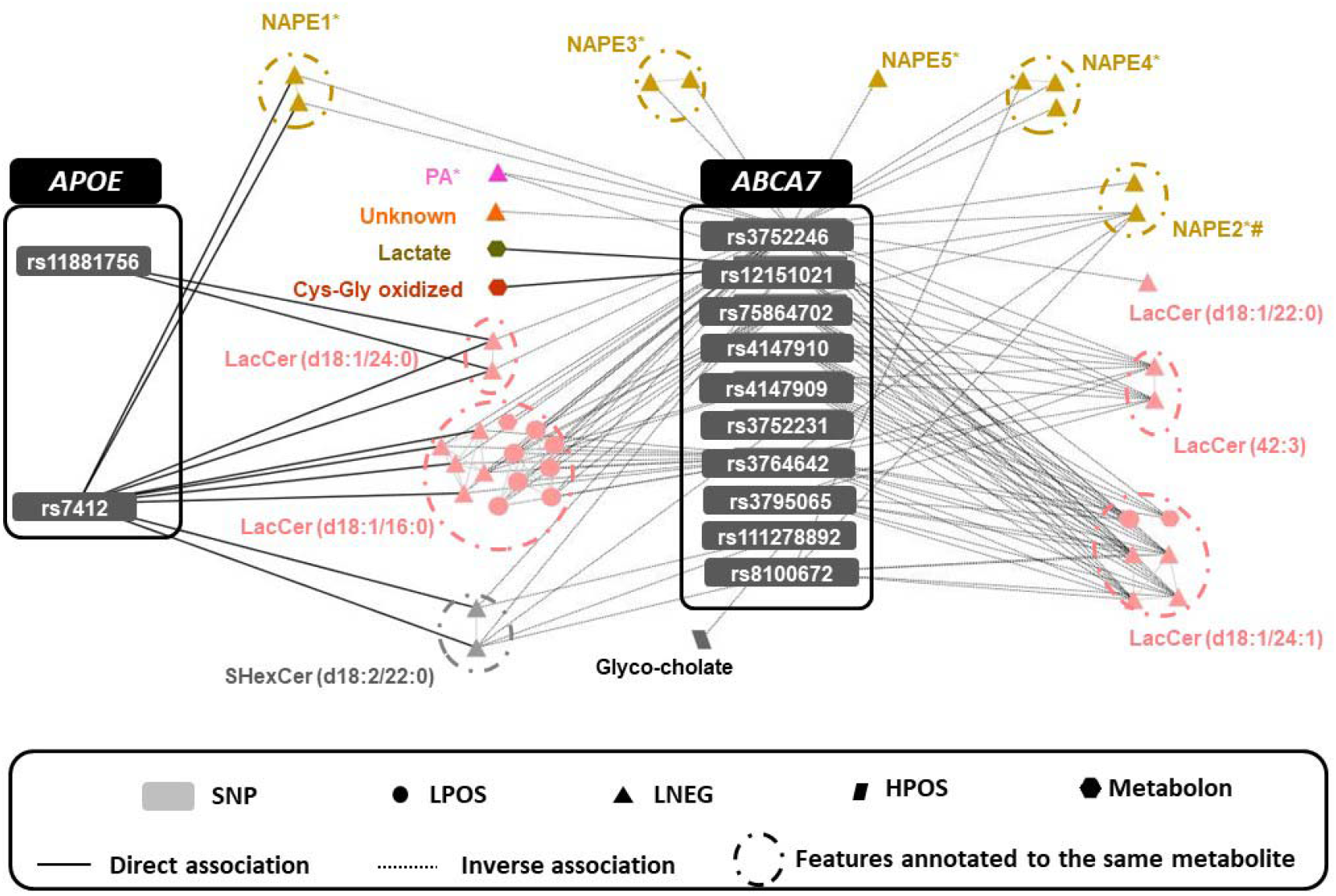
Metabolites associated with AD-related SNPs at *ABCA7* (dark grey) and their association with AD-related SNPs at *APOE* (light grey). A circle surrounds all the multi-platform features annotated to the same metabolite and at least one of those features is associated with the connected SNP. Colours indicate metabolite class and symbols represent the analytical platform used. The features PA*, NAPE4* and NAPE5* contain the carbon side-chain (20:4).

### Metabolomic signatures and cognitive performance

We tested the 14 metabolomic features associated with *ABCA7* detected on the Imperial Phenome Centre platform against individual cognitive performance variation in Airwave and the FINGER trial data using mixed-effect regression models (see <u>Methods</u>). We did not have sufficient number of samples – Airwave only – to test the two metabolites found only on the Metabolon platform. Eleven of these features were annotated in FINGER data (mean age 69.2 ± 4.7 years) and five of these (including three LacCer species) were positively associated (*P* < 0.05/11, Bonferroni threshold) with a composite cognitive score based on an extended version of the Neuropsychological Test Battery^17^. None of these features were associated with cognitive performance among Airwave participants (mean age 40.0 ± 9.2 years), but they were on average more than 20 years younger than FINGER trial participants (**Figure 3-a**). We also tested for associations between changes in these 14 metabolic features in plasma and changes in cognitive scores in the FINGER trial, comparing baseline and two-year follow-up data, and found that higher level of an *N*-acylphosphatidylethanolamine, referred to here as NAPE5* (SLNEG 1015.7305 9.0037), was associated with slower cognitive decline (**Supplementary Figure 3)**.

**Figure 3.**
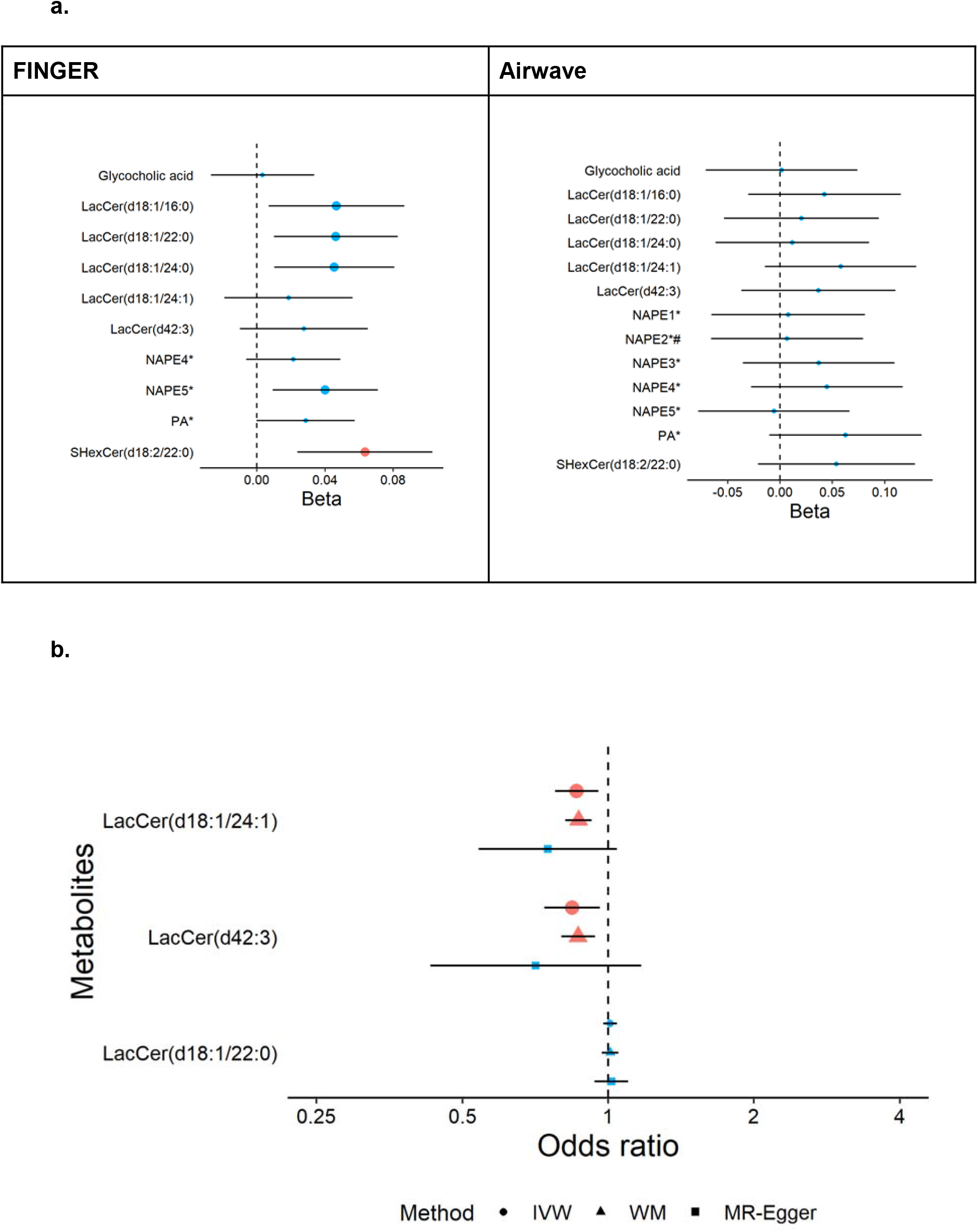
a) The association of *ABCA7*-associated metabolites with cognitive score in FINGER trial and Airwave. For FINGER, the association between metabolite concentrations and cognition was tested using a linear mixed model adjusted for fixed effects of age at baseline, sex, intervention, visit (binary) and domain of participant groups; and for random effects of within-subject variation. For Airwave, the association between metabolite concentrations and cognition at baseline was tested using a linear mixed model adjusted for fixed effects of age at baseline, sex, visit (binary) of participant groups; and for random effects of within-subject variation. We obtained the z-score for both metabolite levels and cognition values prior to analysis. Chemical species are represented by their molecular ion or most abundant adduct ion. Larger dots indicate 95% CI, not including the null value; red dots indicate associations with P<0.05/11 for FINGER and P<0.05/14 for Airwave (Bonferroni correction for 11 and 14 features, respectively). **b) Associations between ABCA7-associated metabolites and risk of AD based on Mendelian randomisation**. Dots indicate inverse variance weighted (IVW) estimates, triangles indicate weighted median (WM) and squares indicate Egger estimates. Red and larger shapes indicate significant associations with P<0.05/3 (Bonferroni correction for 3 features).

### Ceramide metabolism in the brain tissue of the Abca7 knockout mouse

We then carried out metabolic profiling of brain tissue (left hemispheric cerebral cortex) from the *Abca7* KO mouse versus WT to test whether variations in lipid profiles related to *ABCA7* in peripheral blood in the human studies also are found in brain tissue. We performed UPLC-MS in both positive and negative ionisation modes on the lipid phase of chloroform/methanol/water extractions of the cerebral cortex and then performed Principal Component Analysis (PCA) to visualise dominant patterns (**Figure 4**). While the main variance in the positive ionisation mode data was driven by sex (PC1 = 80.4%, PC2 = 8.7 %), the principle factor discriminating samples in the negative ionisation mode data was attributed to genotype (PC1 = 59%, PC2= 14.2%, PC3 = 6.9 %). We also found lower levels of phosphatidylcholines and phosphatidylglycerols in brain tissue from *Abca7* null mice, consistent with previous reports ^14,18^.

**Figure 4.**
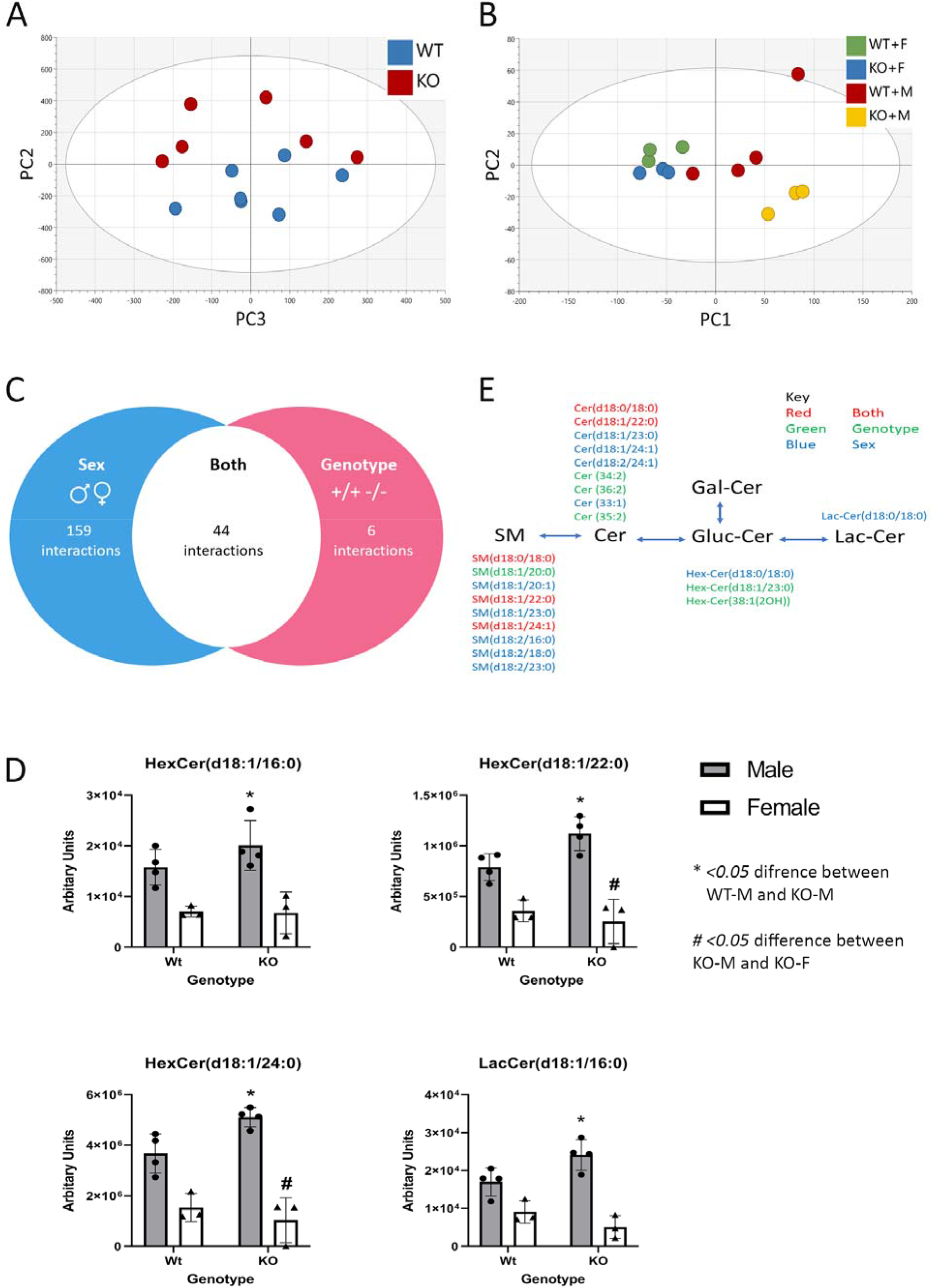
Schematic of the proposed interaction between ceramide metabolism and *ABCA7* in the brain. SNPs associated with loss or reduction in function in *ABCA7* are also associated with AD. *ABCA7* regulates the intracellular lipidome by exporting cholesterol and/or phospholipids out of the cell. We suggest impaired transport via *ABCA7* either directly or indirectly causes an accumulation of intracellular ceramides, hexosylceramide and lactosylceramide. These species are suggested to interact with the cell membrane, altering fluidity and in turn may increase the activity of beta-site *APP* cleaving enzyme to process amyloid precursor protein to produce extracellular amyloid plaques. Alternatively, ceramides may also contribute to increased apoptosis in the brain, similar to pathways detected in peripheral tissue.

Based on associations of plasma LacCer concentrations with the *ABCA7* locus in the human studies, we filtered the lipidomic data specifically for sphingolipids (sphingomyelins, ceramides, and hexose-ceramides) and performed two-way analysis of variance (ANOVA) to investigate genotype-sex interactions. Nine sphingolipids detected in negative mode ionisation differed between *Abca7* KO and WT (*P* < 0.05), with seven associated with genotype and two associated with sex (one passing the Bonferroni correction for multiple testing). Repeating this analysis for the positive mode ionisation dataset, 244 differences were identified across the detected sphingolipids, of which 159 were associated with sex (two passing the Bonferroni correction for multiple testing) and 44 associated with interactions between sex and genotype (24 passing the Bonferroni correction for multiple testing). These data suggest a role for *ABCA7* in regulating ceramide metabolism in the brain, and that may, at least partially, be sex-dependent.

### Mendelian randomisation analysis

Of the 41 mass spectral features related to the 10 AD-related SNPs at *ABCA7*, we annotated eight to three metabolites: LacCer(d18:1/24:1), LacCer(d42:3) and LacCer(d18:1/22:0). From GWAS on Airwave data, we identified genetic instruments (SNPs) for two-sample Mendelian randomisation analysis based on a representative metabolic feature for each of the three metabolites, i.e. LNEG_970.7177_7.8397 for LacCer(d18:1/24:1), LNEG_1066.6794_7.3646 for LacCer(d42:3), and LNEG_1043.6825_7.8568 for LacCer(d18:1/22:0). List of genetic variants for each metabolic feature is presented in **Supplementary Table 3**. Two-sample Mendelian randomisation analysis was performed to assess the association of the metabolic features with risk of AD. Using the inverse variance weighted method, we found supporting evidence for inverse associations of LacCer(d18:1/24:1) and LacCer(d42:3) with AD (**Figure 3-b**). Estimates from weighted median and MR-Egger methods, as sensitivity analyses, were consistent with the inverse variance weighted estimates, although the MR-Egger estimate had wide confidence intervals reflecting its reduced power compared with the other two methods.

## Discussion

We found evidence that LacCers play a role in the association between variants in the *ABCA7* gene and metabolic pathways associated with cognitive performance and AD based on aggregated metabolomic and genomic data from both human and animal studies. Our MWAS indicated that among genes so far identified for AD, *ABCA7* demonstrates the strongest association with metabolic traits in the peripheral blood of humans. We found 121 associations between 10 independent genetic variants at *ABCA7* and mass spectral features in the lipophilic fraction of peripheral blood. Most of the features were annotated to LacCers, with the most robust association being found between rs3752246 and LacCer (d18:1/24:1). Further, we found that deletion of *Abca7* in the mouse results in a range of alterations in sphingolipid metabolism, particularly for ceramides and hexose-ceramides. Moreover, using the Mendelian randomisation approach, we found evidence supporting a potential causal role of LacCers on the risk of AD.

The genetic loci associated with AD can be separated into two broadly-defined groups of genes. The first set are a small number of genes including *APP, PSEN1* and *PSEN2* that have a strong autosomal dominant hereditary effect and associate with familial-early onset AD. For these loci, potential underlying molecular mechanisms have been postulated including that underpinning the influential “amyloid hypothesis” positing that amyloid-beta accumulation results in presenile plaque formation^19-21^. The second group is larger and includes genes such as *APOE* that are associated with sporadic late-onset AD (LOAD). These latter genetic risks interact with environmental factors for the development of AD later in life. Relative to the early onset forms, much less is known about specific mechanisms associated with sporadic LOAD or, indeed, whether there is a common mechanism, or whether a number of distinct mechanistic pathways converge on a common pathology.

*ABCA7* is part of the ATP-binding cassette (ABC) reporter family, important in regulating phospholipid and cholesterol transport across the cell membrane, as well as within the cell between subcellular organelles^18^. *ABCA7* is highly expressed in microglia in cell culture and in neuronal cells, particularly in the hippocampus^22^. From cell culture experiments, *ABCA7* expression is regulated by sterols through the *SREBP2* pathway^23^, and increased expression of *ABCA7* has been shown to enrich HeLa cells with ceramides^24^. Neuronal cells lacking *ABCA7* show evidence for increased ER-stress and Aβ40 and Aβ42 production. Sakae et al. proposed that *ABCA7* deficiency alters lipid metabolism in ways that secondarily increase *SREBP2* and *BACE1* expression and induce ER stress and Aβ accumulation to initiate cognitive deficits that lead to prodromal AD^14^. Consistent with this model, genetic variants at *ABCA7* are associated with relative atrophy of the cortex and hippocampus both in cognitively normal people and those with mild cognitive impairment^25^.

We applied complementary epidemiological resources and approaches to gain new understanding of the pathways linking genetic variants in *ABCA7* to AD risk. Although a large proportion of Airwave^11^ participants are too young to show clinically meaningful cognitive decline, it provides a resource in which relationships between genetic variants and early disturbances in intermediate molecular phenotypes can be explored. We also included data from older adults in FINGER (aged 60 to 77 years at baseline) ^26^ to investigate effects of identified metabolites on cognition. We demonstrated associations of LacCer(d18:1/22:0), LacCer(d18:1/24:0), LacCer(d18:1/16:0), SHexCer, and NAPE5* with cognitive performance. Moreover, we found that levels of NAPE5* were inversely associated with decline in cognitive performance over a two-year period in FINGER (**Supplementary Figure 3**).

Our untargeted lipidomic assay of *Abca7* KO and WT mouse brains showed decreases in a wide range of phosphatidylcholines and phosphatidylglycerols and differences in the concentrations of more than 250 sphingolipids (including ceramides). These results strongly support a role for ABCA7 in sphingolipid/ceramide metabolism. Consistent with this, in an earlier report, Sakae and colleagues used a targeted lipidomics assay of 275 lipids to assess lipidomic changes in the brains of *Abca7* KO mice^14^. They found differences in 24 specific lipids, consisting of decreases in 12 species of phosphatidylethanolamines, three species of phosphatidylglycerols, one lysophosphatidylcholine and two sphingomyelins, and increases in three phosphatidylcholines, one ceramide, three sulfatides and one cerebroside.

We used Mendelian randomisation to test for potential causal relationships between LacCer concentrations in plasma and AD. This is a type of instrumental variable analysis where genetic variants are used as instruments to overcome unmeasured confounding and reverse causation in observational studies ^27^. The two-sample Mendelian randomisation design uses data on risk factors (in this case metabolic features of the LacCers) and outcome (AD) from different cohorts, which allows possibilities to increase statistical power. For our analysis, we first performed a GWAS on metabolic features in Airwave to generate genetic instruments for the target LacCers and then took advantage of the most recent and largest AD GWAS data to test for associations of these genetic variants with AD^2^. Using this approach, we found evidence of a possible causal association of increased concentrations of plasma LacCer (d18:1/24:1) and LacCer (42:3) with decreased risk of AD. Thus, we have found that *ABCA7* contributes to the regulation of systemic LacCer metabolism and that these glycolipids may play causal roles in the genesis of AD.

Gene association, epidemiological studies and animal and cell models all have suggested causal roles for lipid dyshomeostasis in AD pathogenesis, especially for *APOE, ABCA7* and *TREM2*^28^. Gene-set analyses support this by implicating pathways involving lipid metabolism in addition to those for inflammation and amyloid processing^1^. Metabolomic studies of ApoE and Trem2 in mouse models ^28,29^ and cell culture directly implicate lipid dyshomeostasis and inflammation with genetic risk of AD. These observations suggest that the genesis of LOAD may share general features of the metabolic disease-associated inflammation (“meta-inflammation”) that affects adipose and other peripheral tissues in the metabolic syndrome^30^. In this context, we note that four metabolites, NAPE3*, NAPE4*, NAPE5*, PA* and one unassigned metabolite (UA) corresponding to the feature SLNEG_782.4965 3.5812 associated with *ABCA7* SNPs (but not with any of the SNPs at *APOE*) have arachidonic acid (20:4) side chains. The MS/MS experiments for the unassigned metabolite (UA) was also shown to contain a (20:4) acyl chain by tandem mass spectrometry. LacCers activate cPLA2-□, a phospholipase that hydrolyses arachidonic acid from phospholipids^31^. Arachidonic acid, a substrate for cyclooxygenase (COX) and lipoxygenase (LOX), is a driver of macrophage and microglial pro-inflammatory pathways in aging and AD^32^.

Our observations regarding gene-metabolite interactions associated with AD are novel, however, have some limitations. First, we discovered associations of LacCers with the *ABCA7* locus using peripheral blood rather than directly in the brain. Our results thus either imply that systemic biochemical pathology promotes the genesis of AD (e.g., via inflammation) or that there is a related lipid dyshomeostasis in the CNS. However, our observations of differences in concentrations of other lipids (particularly sphingolipids, ceramides, and hexose-ceramides) in the *Abca7* KO mouse brain relative to WT controls are most consistent with the latter possibility. Second, rather than directly testing causal associations between concentrations of LacCers and AD, we carried out Mendelian randomisation analyses in which genetic information was used to identify potential causal links between the LacCers and risk of AD. This approach involves three key assumptions^27^: i) the genetic variants are associated with the risk factor (LacCer metabolites in this case); ii) there are no unmeasured confounders between genetic variants and outcome (AD); and iii) the genetic variants affect the outcome only through their effect on the risk factor (that is, there are no directional pleiotropic effects). We fulfilled the first assumption by setting strict criteria for selecting genetic variants (*P*<5×10^−8^ and F-statistics>10). It is difficult to verify whether the second and third assumptions hold, but our findings were robust to sensitivity analyses and the MR-Egger method did not indicate any potential directional pleiotropic effects.

*ABCA7* is widely expressed throughout the body, including the brain, particularly in myeloid cells (https://www.proteinatlas.org/). AD-risk raising SNPs were all associated with reduced *ABCA7* activity, and Satoh and co-workers have previously shown that knockout of the gene in the mouse increases soluble Aβ in an APP mouse model^33^. From our observations, we propose that in addition to regulating cholesterol and phosphatidylcholine concentrations, *ABCA7* regulates intracellular concentrations of ceramides and hexosyl-ceramides, which were increased in the cortex of *Abca7* KO mice (**Figure 5**). We further hypothesize that the accumulation of these ceramide species in the brain may increase the cleavage of APP to induce plaque formation, as has been previously suggested^14^. This may possibly occur by increasing lipid raft formation, and/or contributing to ER stress, as seen for example in peripheral cell types when exposed to palmitate induced ER-stress through altering both cell membrane composition and dynamics^34^.

**Figure 5.**
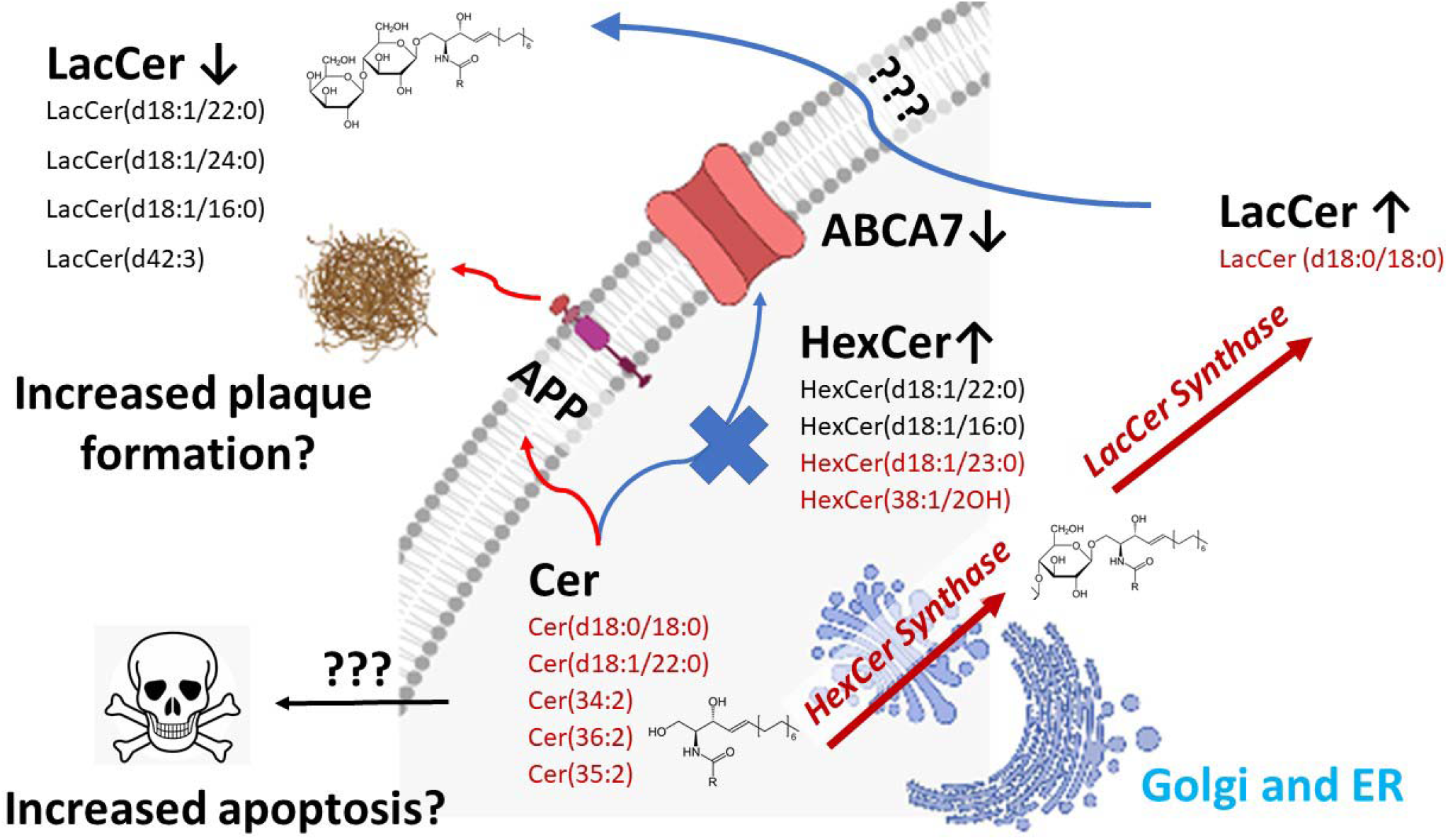
Lipidomics analysis of the cortex of the *Abca7* null mouse identifies alterations in ceramide metabolism in the brain. **A**. Principal components analysis (PCA) of the lipidomic data collected in negative ionisation mode (variation represented by PC2 = 14.2%, PC3 = 6.9%). **B**. PCA of the lipidomic data collected in positive ionisation mode (variation represented by PC1 = 80.4%, PC2 = 8.7%). **C**. Venn diagram of lipidomic changes detected in the dataset summarising all sphingolipid changes associated with genotype (*Abca7*), sex and their interaction by ANOVA. **D**. Box plot of ANOVA results for hexosylceramide (HexCer) (d18:1/16:0), HexCer (d18:1/22:0), and HexCer (d18:1/24:0) and lactosylceramide (LacCer) (d18:1/16:0). **E**. Schematic of key changes detected in sphingomyelins (SM), ceramides (Cer), hexosyl ceramides (HexCer) and lactosyl ceramides (LacCer) detected in both the cortex of the *Abca7* null mouse and the human MWAS analysis. Key: WT wildtype, KO *Abca7* null, m male, f female.

In conclusion, we report associations between SNPs at the *ABCA7* locus and LacCers in peripheral blood, which were associated with differences in cognitive function and potential causal links with risk of AD^35^. Manipulation of sphingolipid metabolism is already being explored for augmenting the efficacy of anti-cancer and insulin resistance therapeutics, effective strategies for which could be tested as “re-purposed” treatments for AD^36^.

## Data Availability

The data is available per request. For cohort data, agreement of the cohort management team is needed.

## Online Methods

### Study samples

Airwave is an occupational cohort of 53,116 police officers and staff ages 18 years and over across Great Britain that was launched in June 2004^11^. Blood samples were obtained at the screening visit (with consent) and stored at −80 °C at the laboratory, before being transferred to the biorepository facility and stored in vapour phase liquid nitrogen. Metabolomic assays were done for approximately 5,250 samples (**Supplementary Table 1**). The samples were divided into two separate sample sets (referred to herein as Airwave 1 and Airwave 2). Airwave 1 utilised lithium heparin plasma samples and Airwave 2 utilised EDTA plasma samples. Both sets of samples were analysed at the Imperial Phenome Centre (London, UK). In addition, a set of 2,250 samples were analysed by Metabolon Inc.TM (Morrisville, NC, USA), of which 1,000 were also included in Airwave 2.

The Rotterdam Study is a prospective cohort study of individuals in the Ommoord district of the city of Rotterdam, the Netherlands^12^. At baseline, between 1990 and 1993, 7,983 participants over 55□years old were interviewed at home and underwent extensive clinical examination at the research centre. Serum samples were collected from 1997 to 1999, stored at −20 °C and analysed for metabolites by Metabometrix Ltd (London, UK) using protocols adopted from the Imperial Phenome Centre. The Airwave and Rotterdam sample and features numbers are described in **Supplementary Table 4**.

FINGER is a randomised controlled trial of 1,260 individuals aged 60–77 years with increased risk for dementia based on the Cardiovascular Risk Factors, Aging, and Incidence of Dementia (CAIDE) dementia risk score and a cognition at mean level or slightly lower than expected for their age^37^. The population was randomly assigned to a multidomain intervention (diet, exercise, cognitive training, vascular risk monitoring) or a control group (general health advice) in a 1:1 ratio. We used metabolomic and cognitive data at baseline and 2-year follow-up. EDTA plasma samples were analysed at the Imperial Phenome Centre.

### Genomics data

For both Airwave and the Rotterdam Study, DNA samples were extracted from leukocytes. Genotyping was done using Illumina Infinium HumanExome-12v1-1 BeadChip Array in Airwave and Illumina 550 K arrays in the Rotterdam Study. Both studies imputed their data to 1000 Genomes Phase 3 reference panel.

### Metabolomics data

For each cohort (Airwave 1, Rotterdam Study and FINGER), metabolomics data were acquired using UPLC-MS. Methods and quality control (QC) have been described previously^38-40^. Briefly, a pooled study reference sample was prepared for each population. Serum and plasma long-term reference samples were prepared using commercial bulk serum and plasma (BioIVT [Seralab], West Sussex, UK). These QC samples were analysed at regular intervals throughout data acquisition. Mixtures of authentic reference standards were added to the study reference, long-term reference, and study samples used in the UPLC-MS analysis to enable targeted monitoring of data quality during acquisition.

Plasma samples from FINGER were prepared and underwent UPLC-MS profiling analysis for lipids and small metabolites as previously described^38^. The same procedures were used for the analysis of plasma samples from Airwave and serum samples from Rotterdam Study except that 100μl of samples was used without dilution prior to addition of isopropanol for the lipidomics analyses. Briefly, batches of 80 samples were prepared into 96-well plates. Each sample was mixed with four parts of 4°C isopropanol, incubated at 4°C, centrifuged, and the supernatant aliquoted into a 96-well plate. All analyses were acquired on Acquity UPLC systems coupled to Xevo G2-S ToF mass spectrometers (Waters Corporation, Milford, Massachusetts, United States). For these samples three platforms were used - reverse phase in positive ionisation mode which detects largely lipid species (Lipid Positive mode - LPOS), reverse phase in negative ionisation mode which detects largely lipid species (Lipid Negative mode - LNEG) and HILIC chromatography in positive mode which detects largely small and polar metabolites (HILIC Positive mode - HPOS).

### Data processing

Peak picking was completed using Bioconductor R-package XCMS^41^. Drift correction was done using a method previously described^42^. For FINGER, we used nPYc toolbox^43^. Negative values were replaced with zeros, and the data were natural log-transformed after adding one. We filtered the data based on retention time to exclude non-retained and cleaning phase features – only features in the following retention times (in minutes) were accepted: HPOS (0.5-7); LNEG (0.3-9.5); LPOS (0.45-12). We used principal components analysis (PCA) to identify samples that were outliers and excluded them. Further, we excluded values that were >5 median absolute deviations (MAD) from the median. We used 10 principal components from genome-wide scans to adjust for population stratification. Finally, we transformed the data into z-scores using median and MAD, so that we had comparable intensities across studies. We determined cross-study feature correspondence for the Airwave and the Rotterdam Study using an in-house algorithm that selects the matches that are the closest in retention time and mass to charge ratio (m/z ratio) in the two pre-aligned datasets.

### UPLC-MS metabolite annotation

Lipid annotation was initially completed by matching accurate mass fragmentation measurements to reference spectra from online databases (LIPID MAPS^44^, Metlin, HMDB) and previous publications. Where chemical reference materials were commercially available (Avanti Polar Lipids, Sigma Aldrich, Cayman Scientific), they were used to generate definitive molecular identification by direct matching of chromatographic and spectral qualities (including accurate mass, MS/MS spectra, and isotopic distribution) to those observed in the profiling data. For FINGER data, annotations were transferred from the Airwave data after finding feature correspondence between the two datasets, using the same method as described above.

### Selection of genetic variants

We identified 47 unique genetic loci based on three recent GWAS on AD^1-3^. The lead SNP from 45 genetic loci were available in our data and was analysed in the exploratory stage MWAS (see below). To focus on the pathways linking *ABCA7* and risk of AD, we selected SNPs with a *P*<1×10^−5^ within +/-500 kb from the lead SNP in *ABCA7* using the most recent GWAS by the International Genomics of Alzheimer’s Project (IGAP) consortium^2^ (Discovery stage: N _case / control_ = 21,982 / 41,944). For correlated SNPs with r^2^≥0.5, we kept the SNP with the smallest *P*-value, resulting in 10 SNPs selected for the MWAS focusing on *ABCA7*.

### Metabolome-wide association study (MWAS)

We carried out a linear regression to study the association of each SNP with all metabolomic features (Metabolon, HPOS, LNEG, LPOS) with adjustment for age, sex, and cohort. We used a permutation-based method to estimate the MWAS Significance Level^45,46^ to account for multiple testing and the high degree of correlation in metabolomics datasets. A *P*-value threshold giving a 5% Family-Wise Error Rate was computed for each SNP in each metabolomics platform.

### Representative features

For subsequent analysis, we used a single feature to represent each metabolite. The representative feature was chosen based on the major detected ion (either parent ion, closest adduct, or isotope which gave the largest intensity and was not confounded by isobaric species).

### Association of metabolites with cognition

We used data from the Airwave and FINGER studies to study the association of identified metabolites with cognitive performance. Cognition was measured using a version of the Cardiff Cognitive Battery^47^ in Airwave and using standard neuropsychological tests (an extended version of the Neuropsychological Test Battery) in FINGER^26^. The analysis for FINGER was based on metabolomics and cognition data both available at baseline and at the follow-up visit. In Airwave, however, the analysis was cross-sectional using the cognition and metabolomic data for individuals at baseline.

### Mendelian randomisation

We performed two-sample Mendelian randomisation analyses to investigate the potential causal relationship between metabolomic features associated with SNPs in *ABCA7* and AD risk. We conducted GWAS on these metabolomic features in Airwave to identify genetic instruments for the Mendelian randomisation analyses. For each feature, data points outside 5 MAD from the median were deleted and subsequently imputed using the k-nearest neighbour (kNN) algorithm (k=10). GWAS were performed using the high-dimensional association analyses (HASE) framework with adjustment for age and sex^48^. Population stratification was controlled by adjusting for 10 principal components during data pre-processing stage. The number of participants analysed in GWAS were 1977 (HPOS), 1979 (LPOS), and 1983 (LNEG).

For each metabolic feature, SNPs with *P*<5×10^−8^ and an F-statistics >10 were selected as genetic instruments for the Mendelian randomisation analysis. To mitigate weak instrument bias, we only included SNPs with a minor allele frequency greater than 5% and imputation quality greater than 0.6. We removed correlated SNPs (r^2^>0.1) by retaining the SNPs with the smallest *P*-value. Mendelian randomisation analyses were performed for metabolic features with more than three independent genetic instruments to allow further sensitivity analyses. The genetic association of AD was based on the most recent GWAS of IGAP^2^.

We searched for genetic instruments for 14 mass spectral features. Three or more independent instruments were available for eight features, annotated to three metabolites. We selected a representative feature for each metabolite for the Mendelian randomisation analysis (**Supplementary Table 4**). We estimated the effect of metabolic features on risk of AD using the inverse variance weighted (IVW) method^49^. Potential pleiotropic effects were assessed using weighted median (WM) and MR-Egger regression as sensitivity analyses. Potential outlier SNPs were identified using MR-PRESSO and were excluded from the analyses^50^. We accounted for multiple testing using Bonferroni correction accounting for the number of tested metabolites (*P<*0.05/3=0.0167).

### Mouse Abca7 knockout model

All animal procedures were approved by the Mayo Clinic’s Institutional Animal Care and Use Committee and were performed in accordance with the National Institutes of Health’s *Guide for the Care and Use of Laboratory Animals^51^*. *Abca7* knockout mice (*Abca7^−/−^*)^52^ were crossbred with WT C57BL/6 inbred mice. Littermate male and female *Abca7^+/+^* (WT) and *Abca7^−/−^* (KO) mice were killed at 50 weeks age (4 WT male, 3 WT female, 4 KO male, 3 KO female, 3 pooled samples) and the left hemisphere of the brain rapidly dissected and frozen on dry ice. Left cerebral cortex (20 mg of wet weight tissue) was extracted using chloroform/methanol/water biphasic extraction and the lipid fraction analysed using UPLC-MS with reverse phase chromatography of the lipid fraction (LPOS and LNEG). Peak picking was performed using the R library XCMS after optimisation of peaks extraction and retention time correction parameters with the library IPO^53^ in conjunction with a tool for automated optimization of XCMS parameters. Further data processing was completed using the KniMet^54^. Briefly, features were removed from further analysis if detected in less than 50% of the pooled samples with a relative LipidSplash internal standard deviation higher than 20%, annotated by exact mass match with the LIPID MAPS database. Missing values were either imputed using the R library “impute”^55^ for multivariate statistical analysis, or replaced with half of the minimum value found in the feature for univariate statistical analysis. In total 10837 features were detected in positive and negative ionisation mode, with 5,503 of these features being assigned by a combination of exact mass, retention time matching and comparison with standards. Multivariate statistics comparing metabolic features detected in KO versus WT mice were performed on both negative and positive ionisation mode data.

## Acknowledgements

This work is supported by the UK Dementia Research Institute at Imperial College, which receives its funding from UK DRI Ltd., funded by the UK Medical Research Council, Alzheimer’s Society and Alzheimer’s Research UK. AD is funded by a Wellcome Trust seed award (206046/Z/17/Z). RM is funded by the President’s PhD Scholarship from Imperial College London. PMM acknowledges generous personal and research support from the Edmond J Safra Foundation and Lily Safra and a National Institute for Health Research (NIHR) Senior Investigator Award (to 2020). JLG is funded by the UK Medical Research Council (MRC) (MC_UP_A090_1006, MC_PC_13030, MR/P011705/1, MR/P01836X/1 and MAP UK) and Wellcome Trust (MetaboFlow). MK is funded by the NIHR Imperial Biomedical Research Centre (BRC), Wallenberg Clinical Scholars, Academy of Finland and Swedish Research Council. AS is funded by the Academy of Finland (287490, 294061, 319318), European Research Council (804371), Alzheimerfonden and Region Stockholm ALF (Sweden). This work was supported by the MRC and NIHR [grant number MC_PC_12025]. PE is director of the MRC Centre for Environment and Health (MR/L01341X/1). He also acknowledges support from the NIHR Health Protection Research Units in Chemical and Radiation Threats and Hazards and in Health Impact of Environmental Hazards. PE is a co-director of the Health Data Research UK London site, which is supported, among others, by MRC, NIHR, Engineering and Physical Sciences Research Council, Economic and Social Research Council, Wellcome Trust, and British Heart Foundation (BHF). PE acknowledges support from the BHF Centre for Research Excellence at Imperial College. Infrastructure support for this research was provided by the NIHR Imperial Biomedical Research Centre (BRC).

## References

1. Jansen, I.E., et al. Genome-wide meta-analysis identifies new loci and functional pathways influencing Alzheimer’s disease risk. Nat Genet 51, 404–413 (2019).

2. Kunkle, B.W., et al. Genetic meta-analysis of diagnosed Alzheimer’s disease identifies new risk loci and implicates Abeta, tau, immunity and lipid processing. Nat Genet 51, 414–430 (2019).

3. Lambert, J.C., et al. Meta-analysis of 74,046 individuals identifies 11 new susceptibility loci for Alzheimer’s disease. Nat Genet 45, 1452–1458 (2013).

4. Lotta, L.A., et al. Genetic Predisposition to an Impaired Metabolism of the Branched-Chain Amino Acids and Risk of Type 2 Diabetes: A Mendelian Randomisation Analysis. PLoS Med 13, e1002179 (2016).

5. Wittemans, L.B.L., et al. Assessing the causal association of glycine with risk of cardio-metabolic diseases. Nat Commun 10, 1060 (2019).

6. Tzoulaki, I., et al. Serum metabolic signatures of coronary and carotid atherosclerosis and subsequent cardiovascular disease. Eur Heart J 40, 2883–2896 (2019).

7. Holmes, E., et al. Human metabolic phenotype diversity and its association with diet and blood pressure. Nature 453, 396–400 (2008).

8. Illig, T., et al. A genome-wide perspective of genetic variation in human metabolism. Nat Genet 42, 137–141 (2010).

9. Nho, K., et al. Altered bile acid profile in mild cognitive impairment and Alzheimer’s disease: Relationship to neuroimaging and CSF biomarkers. Alzheimers Dement 15, 232–244 (2019).

10. Chai, J.F., et al. Associations with metabolites in Chinese suggest new metabolic roles in Alzheimer’s and Parkinson’s diseases. Hum Mol Genet 29, 189–201 (2020).

11. Elliott, P., et al. The Airwave Health Monitoring Study of police officers and staff in Great Britain: rationale, design and methods. Environ Res 134, 280–285 (2014).

12. Ikram, M.A., et al. Objectives, design and main findings until 2020 from the Rotterdam Study. Eur J Epidemiol 35, 483–517 (2020).

13. Marengoni, A., et al. The Effect of a 2-Year Intervention Consisting of Diet, Physical Exercise, Cognitive Training, and Monitoring of Vascular Risk on Chronic Morbidity-the FINGER Randomized Controlled Trial. J Am Med Dir Assoc 19, 355–360 e351 (2018).

14. Sakae, N., et al. ABCA7 Deficiency Accelerates Amyloid-beta Generation and Alzheimer’s Neuronal Pathology. J Neurosci 36, 3848–3859 (2016).

15. Smith, G.D. & Ebrahim, S. ‘Mendelian randomization’: can genetic epidemiology contribute to understanding environmental determinants of disease? Int J Epidemiol 32, 1–22 (2003).

16. Consortium, G.T. The GTEx Consortium atlas of genetic regulatory effects across human tissues. Science 369, 1318–1330 (2020).

17. Harrison, J., et al. A neuropsychological test battery for use in Alzheimer disease clinical trials. Arch Neurol 64, 1323–1329 (2007).

18. Aikawa, T., Holm, M.L. & Kanekiyo, T. ABCA7 and Pathogenic Pathways of Alzheimer’s Disease. Brain Sci 8(2018).

19. Gotz, J., Chen, F., van Dorpe, J. & Nitsch, R.M. Formation of neurofibrillary tangles in P301l tau transgenic mice induced by Abeta 42 fibrils. Science 293, 1491–1495 (2001).

20. LaFerla, F.M., Tinkle, B.T., Bieberich, C.J., Haudenschild, C.C. & Jay, G. The Alzheimer’s A beta peptide induces neurodegeneration and apoptotic cell death in transgenic mice. Nat Genet 9, 21–30 (1995).

21. Oddo, S., et al. Triple-transgenic model of Alzheimer’s disease with plaques and tangles: intracellular Abeta and synaptic dysfunction. Neuron 39, 409–421 (2003).

22. Kim, W.S., Guillemin, G.J., Glaros, E.N., Lim, C.K. & Garner, B. Quantitation of ATP-binding cassette subfamily-A transporter gene expression in primary human brain cells. Neuroreport 17, 891–896 (2006).

23. Iwamoto, N., Abe-Dohmae, S., Sato, R. & Yokoyama, S. ABCA7 expression is regulated by cellular cholesterol through the SREBP2 pathway and associated with phagocytosis. J Lipid Res 47, 1915–1927 (2006).

24. Kielar, D., et al. Adenosine triphosphate binding cassette (ABC) transporters are expressed and regulated during terminal keratinocyte differentiation: a potential role for ABCA7 in epidermal lipid reorganization. J Invest Dermatol 121, 465–474 (2003).

25. Ramirez, L.M., et al. Common variants in ABCA7 and MS4A6A are associated with cortical and hippocampal atrophy. Neurobiol Aging 39, 82–89 (2016).

26. Ngandu, T., et al. A 2 year multidomain intervention of diet, exercise, cognitive training, and vascular risk monitoring versus control to prevent cognitive decline in at-risk elderly people (FINGER): a randomised controlled trial. Lancet 385, 2255–2263 (2015).

27. Davies, N.M., Holmes, M.V. & Davey Smith, G. Reading Mendelian randomisation studies: a guide, glossary, and checklist for clinicians. BMJ 362, k601 (2018).

28. Nugent, A.A., et al. TREM2 Regulates Microglial Cholesterol Metabolism upon Chronic Phagocytic Challenge. Neuron 105, 837–854 e839 (2020).

29. Zhao, N., et al. Alzheimer’s Risk Factors Age, APOE Genotype, and Sex Drive Distinct Molecular Pathways. Neuron 106, 727–742 e726 (2020).

30. Li, C., et al. Macrophage polarization and meta-inflammation. Transl Res 191, 29–44 (2018).

31. Nakamura, H. & Murayama, T. Role of sphingolipids in arachidonic acid metabolism. J Pharmacol Sci 124, 307–312 (2014).

32. Wang, P., et al. Aggravation of Alzheimer’s disease due to the COX-2-mediated reciprocal regulation of IL-1beta and Abeta between glial and neuron cells. Aging Cell 13, 605–615 (2014).

33. Satoh, K., Abe-Dohmae, S., Yokoyama, S., St George-Hyslop, P. & Fraser, P.E. ATP-binding cassette transporter A7 (ABCA7) loss of function alters Alzheimer amyloid processing. J Biol Chem 290, 24152–24165 (2015).

34. Han, J. & Kaufman, R.J. The role of ER stress in lipid metabolism and lipotoxicity. J Lipid Res 57, 1329–1338 (2016).

35. Lewis, A.C., Wallington-Beddoe, C.T., Powell, J.A. & Pitson, S.M. Targeting sphingolipid metabolism as an approach for combination therapies in haematological malignancies. Cell Death Discov 4, 72 (2018).

36. Chaurasia, B., et al. Targeting a ceramide double bond improves insulin resistance and hepatic steatosis. Science 365, 386–392 (2019).

37. Kivipelto, M., et al. The Finnish Geriatric Intervention Study to Prevent Cognitive Impairment and Disability (FINGER): study design and progress. Alzheimers Dement 9, 657–665 (2013).

38. Izzi-Engbeaya, C., et al. The effects of kisspeptin on beta-cell function, serum metabolites and appetite in humans. Diabetes Obes Metab 20, 2800–2810 (2018).

39. Lewis, M.R., et al. Development and Application of Ultra-Performance Liquid Chromatography-TOF MS for Precision Large Scale Urinary Metabolic Phenotyping. Anal Chem 88, 9004–9013 (2016).

40. Dona, A.C., et al. Precision high-throughput proton NMR spectroscopy of human urine, serum, and plasma for large-scale metabolic phenotyping. Anal Chem 86, 9887–9894 (2014).

41. Smith, C.A., Want, E.J., O’Maille, G., Abagyan, R. & Siuzdak, G. XCMS: processing mass spectrometry data for metabolite profiling using nonlinear peak alignment, matching, and identification. Anal Chem 78, 779–787 (2006).

42. Dunn, W.B., et al. Procedures for large-scale metabolic profiling of serum and plasma using gas chromatography and liquid chromatography coupled to mass spectrometry. Nat Protoc 6, 1060–1083 (2011).

43. Sands, C.J., et al. The nPYc-Toolbox, a Python module for the pre-processing, quality-control and analysis of metabolic profiling datasets. Bioinformatics 35, 5359–5360 (2019).

44. Fahy, E., Sud, M., Cotter, D. & Subramaniam, S. LIPID MAPS online tools for lipid research. Nucleic Acids Res 35, W606–612 (2007).

45. Chadeau-Hyam, M., et al. Metabolic profiling and the metabolome-wide association study: significance level for biomarker identification. J Proteome Res 9, 4620–4627 (2010).

46. Castagne, R., et al. Improving Visualization and Interpretation of Metabolome-Wide Association Studies: An Application in a Population-Based Cohort Using Untargeted (1)H NMR Metabolic Profiling. J Proteome Res 16, 3623–3633 (2017).

47. Gallacher, J., et al. A platform for the remote conduct of gene-environment interaction studies. PLoS One 8, e54331 (2013).

48. Roshchupkin, G.V., et al. HASE: Framework for efficient high-dimensional association analyses. Sci Rep 6, 36076 (2016).

49. Burgess, S. & Thompson, S.G. Interpreting findings from Mendelian randomization using the MR-Egger method. Eur J Epidemiol 32, 377–389 (2017).

50. Verbanck, M., Chen, C.Y., Neale, B. & Do, R. Detection of widespread horizontal pleiotropy in causal relationships inferred from Mendelian randomization between complex traits and diseases. Nat Genet 50, 693–698 (2018).

51. National Research Council (U.S.). Committee for the Update of the Guide for the Care and Use of Laboratory Animals., Institute for Laboratory Animal Research (U.S.) & National Academies Press (U.S.). Guide for the care and use of laboratory animals. xxv, 220 p (National Academies Press,, Washington, D.C., 2011).

52. Kim, W.S., et al. Abca7 null mice retain normal macrophage phosphatidylcholine and cholesterol efflux activity despite alterations in adipose mass and serum cholesterol levels. J Biol Chem 280, 3989–3995 (2005).

53. Libiseller, G., et al. IPO: a tool for automated optimization of XCMS parameters. BMC Bioinformatics 16, 118 (2015).

54. Liggi, S., et al. KniMet: a pipeline for the processing of chromatography-mass spectrometry metabolomics data. Metabolomics 14, 52 (2018).

55. Hastie, T.T. R; Narasimhan, B; Chu, G. impute: impute: Imputation for microarray data. (2016).

