## Supplementary document for "Metabolome-wide association study on *ABCA7* demonstrates a role for ceramide metabolism in impaired cognitive performance and Alzheimer’s disease"

**Supplementary Figure legends**

**Supplementary Figure 1** – **Overall design of the study**

**Supplementary Figure 2**. **Expression quantitative trait locus (eQTL) of ABCA7-related SNPs in whole blood, liver, and brain tissues** (* False discovery rate<0.05 ; ** P<0.05/27).

**Supplementary Figure 3. The association of change in metabolite levels with change in cognitive score in FINGER trial**

**Supplementary Table 1** - Number of data points, metabolites and metabolomic features associated with selected SNPs. Key: Metabolon: performed using the Metabolon platform. LPOS: Reverse phase chromatography, positive ionisation mode mass spectrometry. LNEG: Reverse phase chromatography, negative ionisation mode mass spectrometry. HPOS: HILIC positive chromatography, positive ionisation mode mass spectrometry (Imperial Phenome Centre)

| **Assay** | **Airwave study**  **Number of samples** | **Rotterdam Study**  **Number of samples** | **Detected features / annotated features for each platform** | **Number of features associated with all AD-related SNPs** | **Significance threshold for association with SNPs at *ABCA7*** | **Number of features /metabolites associated with SNPs at *ABCA7*** |
| --- | --- | --- | --- | --- | --- | --- |
| LPOS | 1979 | 555 | 1848 / 644 | 29 | 6.0x10^-5^ - 6.7x10^-5^ | 8/2 |
| LNEG | 1983 | 514 | 1460 / 146 | 42 | 5.4x10^-5^ - 6.3x10^-5^ | 28/13 |
| HPOS | 1977 | 524 | 748 / 196 | 3 | 1.0x10^-4^ | 1/1 |
| Metabolon Inc.™ | 2250 | NA | 1143 / 829 | 12 | 5.9x10^-5^ - 6.6x10^-5^ | 4/4 |

**Supplementary Table 2** - MWAS on 10 *ABCA7* SNPs using untargeted MS platforms (LNEG, LPOS, HPOS) as well as Metabolon

Attached as an excel file

**Supplementary Table 3 -** Genetic instruments used for Mendelian randomisation analysis on *ABCA7*-related metabolic features

| **Feature*** | **Annotation** | **SNP** | **EA** | **OA** | **EAF** | **Beta** | **SE** | **P-value** | **FS** | **PVE** |
| --- | --- | --- | --- | --- | --- | --- | --- | --- | --- | --- |
| PLNEG_970.7177_7.8397 | LacCer(d18:1/24:1) | rs4147929 | A | G | 0.18 | -0.654 | 0.051 | 4.20E-38 | 167 | 0.08 |
| PLNEG_970.7177_7.8397 | LacCer(d18:1/24:1) | rs4147904 | A | G | 0.49 | -0.346 | 0.04 | 8.30E-18 | 74 | 0.04 |
| PLNEG_970.7177_7.8397 | LacCer(d18:1/24:1) | rs3761026 | T | C | 0.58 | -0.337 | 0.046 | 2.60E-13 | 54 | 0.03 |
| PLNEG_970.7177_7.8397 | LacCer(d18:1/24:1) | rs73503267 | A | G | 0.45 | -0.265 | 0.043 | 6.40E-10 | 38 | 0.02 |
| PLNEG_1066.6794_7.3646 | LacCer(d42:3) | rs4147929 | A | G | 0.18 | -0.532 | 0.051 | 1.50E-25 | 109 | 0.05 |
| PLNEG_1066.6794_7.3646 | LacCer(d42:3) | rs4147904 | A | G | 0.49 | -0.279 | 0.04 | 3.90E-12 | 48 | 0.02 |
| PLNEG_1066.6794_7.3646 | LacCer(d42:3) | rs3761026 | T | C | 0.58 | -0.294 | 0.046 | 1.20E-10 | 41 | 0.02 |
| PLNEG_1066.6794_7.3646 | LacCer(d42:3) | rs73503267 | A | G | 0.45 | -0.255 | 0.043 | 2.00E-09 | 36 | 0.02 |
| PLNEG_1043.6825_7.8568 | LacCer(d18:1/22:0) | rs78018648 | C | T | 0.91 | 0.886 | 0.069 | 1.40E-37 | 164 | 0.08 |
| PLNEG_1043.6825_7.8568 | LacCer(d18:1/22:0) | rs2071846 | G | T | 0.87 | 0.574 | 0.072 | 2.30E-15 | 63 | 0.03 |
| PLNEG_1043.6825_7.8568 | LacCer(d18:1/22:0) | rs5758865 | G | T | 0.61 | -0.281 | 0.041 | 3.90E-12 | 48 | 0.02 |
| PLNEG_1043.6825_7.8568 | LacCer(d18:1/22:0) | rs10460588 | C | T | 0.81 | -0.313 | 0.049 | 2.00E-10 | 40 | 0.02 |
| PLNEG_1043.6825_7.8568 | LacCer(d18:1/22:0) | rs78733497 | C | T | 0.93 | 0.466 | 0.079 | 3.80E-09 | 35 | 0.02 |
| PLNEG_1043.6825_7.8568 | LacCer(d18:1/22:0) | rs5996230 | A | G | 0.62 | 0.242 | 0.041 | 4.10E-09 | 35 | 0.02 |
| PLNEG_1043.6825_7.8568 | LacCer(d18:1/22:0) | rs2366959 | C | T | 0.11 | -0.369 | 0.063 | 4.90E-09 | 34 | 0.02 |
| PLNEG_1043.6825_7.8568 | LacCer(d18:1/22:0) | rs6519337 | C | T | 0.7 | 0.256 | 0.044 | 6.30E-09 | 34 | 0.02 |
| PLNEG_1043.6825_7.8568 | LacCer(d18:1/22:0) | rs174541 | T | C | 0.65 | 0.234 | 0.042 | 2.20E-08 | 31 | 0.02 |
| PLNEG_1043.6825_7.8568 | LacCer(d18:1/22:0) | rs6002756 | C | T | 0.49 | -0.234 | 0.042 | 3.30E-08 | 31 | 0.02 |

* platform_m/z ratio_retension time

SNP (single nucleotide polymorphism), EA (Effective allele), OA (Other allele), EAF (effect allele frequency), SE (standard error), FS (F-statistics), PVE (proportion of variance explained)

**Supplementary Figure1.**

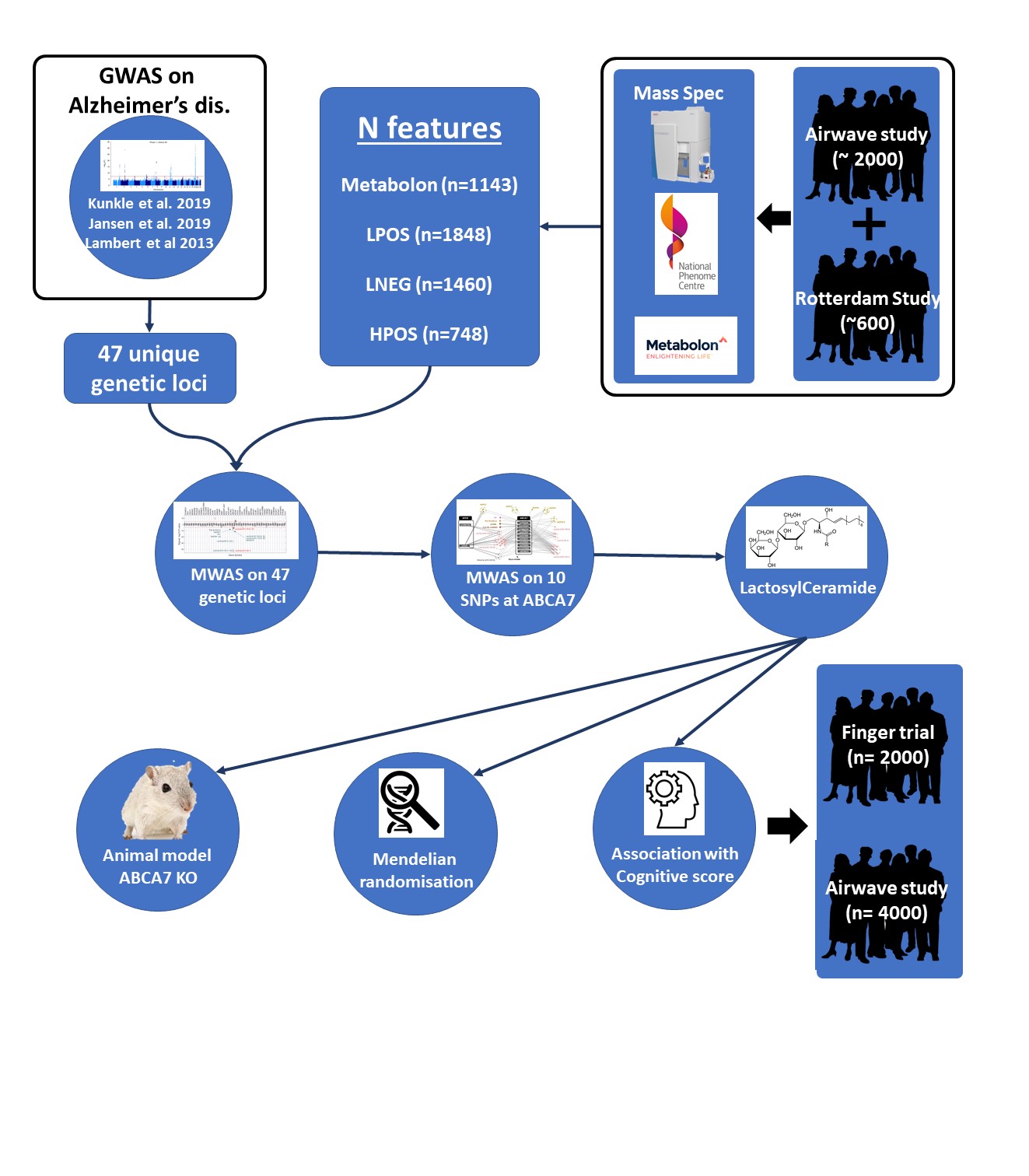

**Supplementary Figure 2.**

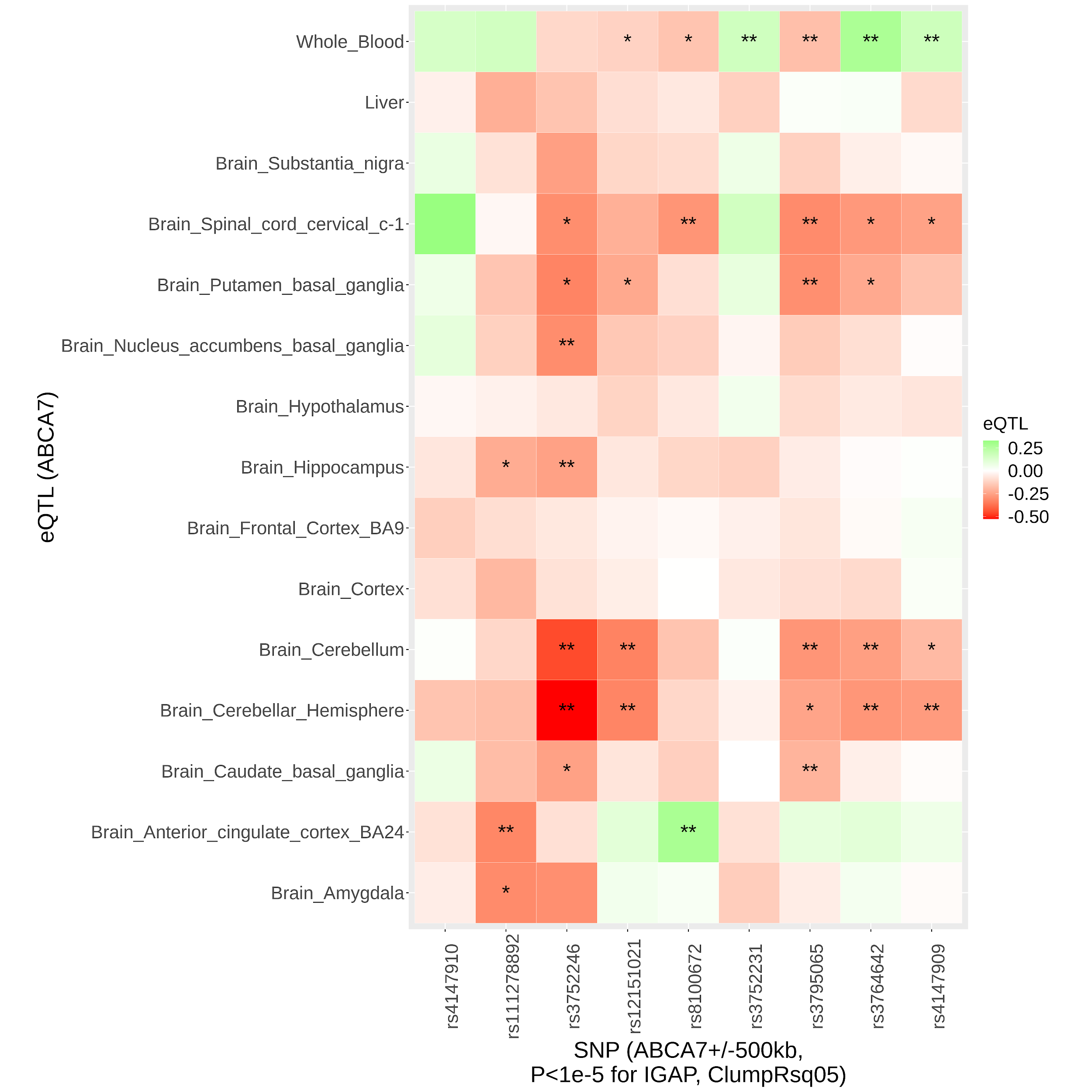

**Supplementary Figure 3.**

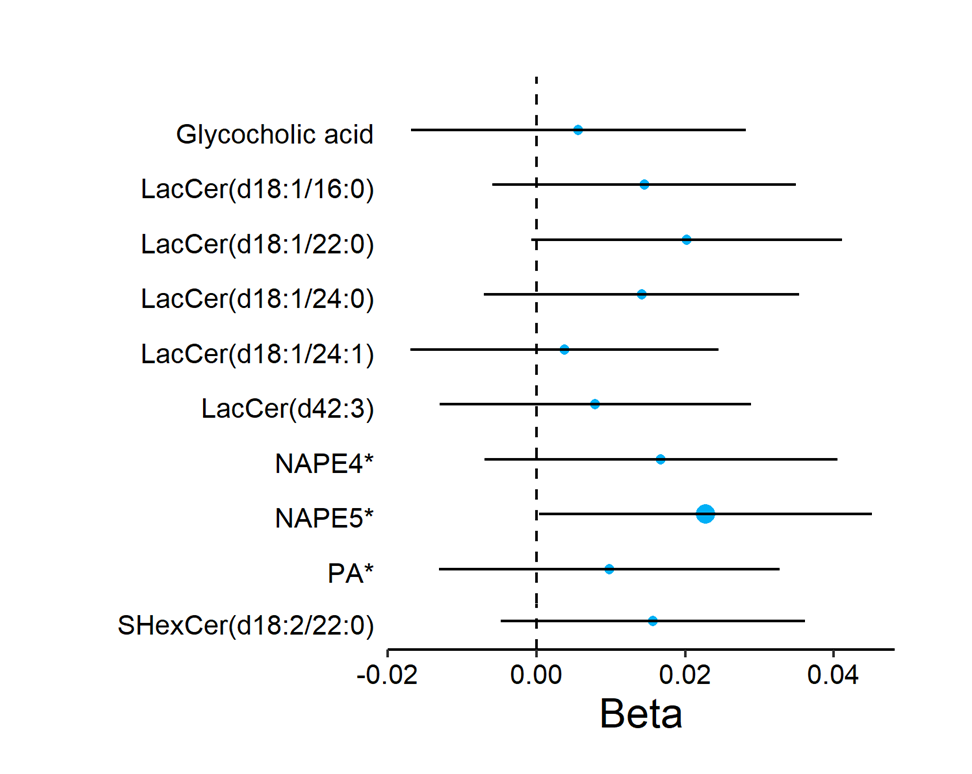
